# Thalamic organization in essential tremor patients with neuropathy: implication for functional neurosurgery

**DOI:** 10.1101/2021.02.21.21251847

**Authors:** Francesco Sammartino, Vinayak Narayan, Barbara Changizi, Aristide Merola, Vibhor Krishna

## Abstract

**Background:** Mechanisms underlying the suboptimal effect of ventral intermediate nucleus deep brain stimulation in patients with essential tremor and co-morbid peripheral neuropathy remain unclear.

**Objectives:** We compared disease-related (location and extension of the ventral intermediate nucleus) and surgery-related (targeting, intraoperative testing) factors in essential tremor patients with and without peripheral polyneuropathy treated with deep brain stimulation of the ventral intermediate nucleus, testing whether the overlap between volume of tissue activated and ventral intermediate nucleus (target coverage) was associated with clinical outcomes.

**Methods:** Preoperative diffusion magnetic resonance imaging was used for thalamic segmentation, based on preferential cortical connectivity. The target coverage was estimated using a finite element model. Tremor severity was scored at rest, posture, action, and handwriting at baseline, 6, and 12 months. Tremor improvement <50% at 12 months was deemed suboptimal. Vertex-wise shape analysis and edge analysis were performed to compare the ventral intermediate nucleus location and extension.

**Results:** 9.7% (18/185) of essential tremor patients treated with deep brain stimulation had co-morbid polyneuropathy. These patients showed a more medial (p=0.03) and anterior (p=0.04) location of the ventral intermediate nucleus, lower target coverage (p=0.049), and worse clinical outcomes (p=0.006) compared to those without polyneuropathy. No differences were observed in the volume of tissue activated between the two groups. Optimal clinical outcomes were associated with greater target coverage (optimal coverage >48%).

**Conclusions:** In essential tremor, co-morbid polyneuropathy may result in suboptimal deep brain stimulation outcomes and lower target coverage, likely related to a reorganization of the ventral thalamic nuclei.

## Introduction

With a prevalence of 4% in the adult population ^1^, essential tremor (ET) is the most common movement disorder and a leading cause of functional and psychological disabilities ^2, 3^. Critically, no medications are available for ET apart from beta-blockers and sedatives (i.e., benzodiazepines, barbiturates, gabapentin), which have a trade-off between symptomatic tremor control and side effects such as drowsiness, fatigue, and confusion ^4-6^. Consequently, over 20% of ET patients cannot achieve reasonable tremor control with medications and require considering surgical treatments.

Deep brain stimulation (DBS) of the ventral intermediate thalamic nucleus (VIM) is a well-established interventional treatment for ET with clinical results significantly superior to oral medications alone ^7, 8^. Still, 10-20% of VIM-DBS cases may report incomplete tremor control ^9, 10^, attributed to inaccurate lead placement ^11^, advanced age, high tremor severity scores ^12^, and development of tolerance to VIM-DBS ^13^. Co-morbid polyneuropathy (PNP), a frequent ET comorbidity affecting ∼10% of the population between 60 and 80 ^14^, may also adversely impact the VIM-DBS outcomes ^15-17^. However, fundamental pathogenic mechanisms of DBS failure in ET patients with PNP remain unclear. A recent review classified potential causes of suboptimal outcomes after VIM-DBS into disease-related, surgery-related, and programming-related factors ^18^. Advanced programming options such as cycling were shown to salvage early (within one year of implantation) DBS failure in neuropathy patients^16^. However, additional mechanisms outside of programming-related factors remain unclear.

We sought to investigate differences in disease-related (VIM extension and location) and surgery-related (stereotactic coordinates, number of passes, VIM length, sensory threshold) factors in ET patients with and without PNP. We also aimed to test whether the convergence of disease- and surgery-related factors, as measured by the overlap of the volume of tissue activated with the ventral intermediate nucleus (target coverage) was associated with tremor outcomes. We used the diffusion magnetic resonance imaging (dMRI) connectivity analysis to compare the VIM location and extension in patients with and without PNP, analyze the overlap between VIM and the volume of tissue activated (VTA) from DBS programming, and compare the outcomes of VIM-DBS in patients with and without PNP at 12-months.

## Methods

### Study Population

We searched the clinical and imaging registry of patients treated with VIM-DBS at The Ohio State University (OSU) Center for Neuromodulation, which includes a vast repository of over 400 cases of movement disorder patients treated with DBS between 2010 and 2019, focused ultrasound ablation, and other interventional therapies. All patients provided written informed consent to be included in this registry, and the institutional review board approved the study.

*Inclusion criteria* were (1) PNP, defined as the bilateral, length-dependent reduction in discriminatory tactile, proprioceptive, or vibratory sensory ability, tendon reflexes, or motor functions ^19^ supported by a reduction in the nerve conduction velocity or action potential amplitude; (2) ET, defined as a bilateral action and posture tremor in 4-10 Hz frequency along a defined axis, without sudden jerky movements or sudden changes in amplitude in the absence of bradykinesia, rigidity, rest tremor, and gait disturbance ^20^; (3) VIM-DBS, conducted at OSU after multidisciplinary evaluation; and (4) Post-surgical follow-up duration of at least 12 months.

*Exclusion criteria* were (1) surgical complication (e.g., stroke or infection) potentially influencing the clinical outcomes; (2) incomplete clinical or imaging data; (3) tremor disorders other than ET. In particular, the differential diagnosis of ET associated with PNP (ET/N+) vs. neuropathic tremor was based on an accurate diagnostic screening including the following clinical and phenomenological factors: frequency and amplitude of tremor in proximal and distal districts, family history of tremor, evidence of inflammatory peripheral neuropathy meeting the criteria for chronic inflammatory demyelinating neuropathy (CIDP), multifocal motor neuropathy (MMN), and IgM demyelinating paraprotein-related neuropathy ^21^.

After identifying ET/N+ patients meeting the criteria listed above, an equal number of age-(±5 years) and gender-matched controls without PNP (ET/N-) was consecutively selected from the same database.

### Surgical procedure and post-surgical DBS programming

The surgical procedure of VIM-DBS was carried out using intraoperative recordings and testing. The VIM was targeted using a combination of formulaic methods and further refined with patient-specific anatomical details such as the size of the third ventricle, the distance between the anterior and posterior commissures, and tractography-based information ^22, 23^.

Local anesthesia and intravenous sedation were administered for burr-hole placement, following which microelectrode recordings (MER) and macrostimulation were performed. The following intraoperative findings verified the electrode location in the VIM: (1) presence of neurons responsive to movements and neurons firing at frequency time-locked with the patient’s tremor, (2) tremor arrest during macroelectrode stimulation. The electrode positioning was optimized for a clinically acceptable therapeutic window (amplitude threshold for side effect minus amplitude threshold for tremor reduction, 1.5 milliamperes or higher) by determining the thresholds for ataxia (indicating cephalon-caudal location relative to the ventral thalamic border), motor contractions (indicating mediolateral location relative to the pyramidal tract), and persistent paresthesia (indicating anteroposterior location relative to the medial lemniscus). Once the localization was verified with physiology, the DBS lead was inserted to the desired depth under fluoroscopic guidance. All patients underwent a postoperative CT scan (voxel size: (voxel size 0.51×0.51×1mm) to determine electrode location and placement accuracy.

Post-surgical DBS programming was based on a monopolar review of each contact of the DBS electrode. The contacts with the best tremor reduction and therapeutic window were subsequently used for stimulation, prioritizing contacts with the most efficient stimulation delivery (largest therapeutic window and least amplitude threshold for tremor reduction). In the case of a suboptimal therapeutic window resulting in incomplete tremor control or side effects limiting the possibility of increasing the stimulation amplitude to the therapeutic levels, selected bipolar configurations were tested to find the stimulation setting providing the best clinical results.

### Imaging analysis

#### Acquisition

Preoperative diffusion MRI was obtained using a 3 Tesla MRI with 3D T1 (1mm isovoxel, TFE) and diffusion-weighted imaging (64 directions, 2mm isovoxel, 71 axial slices covering the whole brain; diffusion-weighting with b = 1000 s/mm2 was applied along 61 directions, uniformly distributed on the sphere, and one b = 0s volume was also acquired) using a padded 32-channel birdcage coil to minimize discomfort and head motion.

#### Pre-processing

Diffusion MRI data were processed with FSL (FSL, Oxford, UK) after conversion to NifTi format. Motion and EDDY artifacts were corrected using EDDY implementation in FSL. BEDPOSTX was used for Bayesian estimation of the diffusion parameters in each voxel (3 fibers model). We calculated a nonlinear transform from diffusion to ICBM template space for each patient’s T1 image, using FLIRT and FNIRT algorithms in FSL. Using the DTIFIT software, we estimated voxel-wise diffusion tensor (DT) for further analysis.

#### Thalamic parcellation using probabilistic tractography

The thalamus mask derived from the Harvard-Oxford subcortical atlas was used to segment the thalamus. To ensure all thalamic voxels across subjects were incorporated, the thalamic mask was thresholded to 70% of the initial segmentation. We then seeded voxels in the thalamus to prespecified cortical areas (motor cortex, premotor & supplementary motor, sensory, prefrontal, temporal, posterior parietal, and occipital cortex) as end regions using a methodology previously described.^24^ Briefly, each thalamic voxels’ connectivity distribution to the seven cortical areas was obtained using Probtrackx2 software (FSL, Oxford, UK). For this analysis, we used 5000 samples per voxel, curvature threshold of 0.2, the maximum number of steps per sample of 2000, step length of 0.5 mm, subsidiary fiber volume threshold of 0.01, loopback check, and no exclusion mask. A hard segmentation of the thalamus was then performed by classifying the seed voxel as connecting to the cortical mask with the highest connection probability. This methodology resulted in the following three parcels within the ventrolateral thalamus from anterior to posterior: ventral oralis anterior & posterior (VOA/VOP), VIM ventro-caudalis sensory nucleus (VC).

### Calculation of the Volume of Tissue Activated (VTA)

LeadDBS, an open-source software ^25^, was used to determine DBS contacts’ spatial location in the patient space by co-registering pre-(T1-weighted MR images) and postoperative (computed tomographic) imaging.^25^ The images were then normalized and co-registered to the 0.5 mm ICBM template using the ANTs implementation. We generated individual VTA models for each patient using DBS settings at initial and 12-months follow-up programming. The calculation of stimulation volumes was based on finite element modeling by accounting for the differences between gray matter, white matter, and ventricle locations. The VTA at 12 months was overlapped with the VIM nucleus to calculate the percentage overlap for each patient using AfNI (NIH, Bethesda, MD). The stimulation volumes were grouped and threshold to 75% of the overlap to define the grouped stimulation volume similar to the methodology described previously.^26^

### Clinical data

Clinical data were collected at the following time points: 1) preoperative baseline, 2) six-month post-surgical follow-up, and 3) 12 months post-surgical follow-up. Baseline data included age, gender, handedness, disease duration, family history of tremor, alcohol responsiveness, the underlying cause for polyneuropathy, and the Fahn-Tolosa-Marin Tremor Rating Scale (TRS) in the treated hand at rest, posture, action, and during handwriting^27^. A detailed registry of the medications for tremor (beta-blockers, primidone, benzodiazepines (BDZ), topiramate, and gabapentin) was collected at baseline and follow-up. The DBS settings at initial and final programming visits were recorded.

## Statistical analysis

The volume, shape, and location of the VIM, VOA/VOP, and VC parcels with vertex-wise shape analysis were compared using t-tests with 3dclustSim software using correction of multiple comparisons (AFNI, NIH, Bethesda, MD) based on a methodology previously reported ^28^. For this analysis, we used 10000 random permutations and a cluster corrected threshold corresponding to a p-value=0.049. Additionally, we compared the extension of each thalamic parcel between ET/N+ vs. ET/N- by calculating the difference in the coordinates of the edges found to be significantly different by the shape analysis.

The following surgery-related factors were compared between the ET/N+ and ET/N- cohorts: stereotactic coordinates, number of passes required, length of the VIM determined by intraoperative physiology, and threshold for persistent paresthesia.

Percentage tremor improvement was calculated by subtracting the 12-month TRS from baseline values. Suboptimal outcomes were defined as a tremor improvement < 50%. A chi-square or t-test was used to compare categorical and continuous variables, as appropriate.

A recursive partitioning analysis (RPA) was used to build decision trees and model predictors, and prognostic groups using the statistical package rpart in R ^29^. The primary outcome variable was dichotomized based on the presence or absence of 50% or greater tremor improvement after VIM DBS. The predictive variables included patient cohort (ET/N+ vs. ET/N-), percentage overlap of the stimulation volume at 12 months with the VIM (continuous variable). Each variable was examined for the best within variable and population-level split. The node was split if the modified Wilcoxon statistic was significant for any variable beyond the .05 probability level ^30^. The coordinates of the center of mass and edges were calculated separately in three axes of space for ET/N+ and ET/N-. A difference of more than 2 mm in either of the three axes was considered significant.

Data were expressed as mean ± standard deviation (SD) unless specified differently. A p-value of less than 0.05 was considered significant. The statistical analysis was performed in R (R 3.5.3 5; R Foundation for Statistical Computing).

## Results

### Clinical and Demographics Data

The searching criteria resulted in n=18 ET/N+ patients (4 females), corresponding to 9.7% of the entire ET registry dataset. Eighteen age- and sex-matched ET/N- controls were included in the analyses. No differences were observed in the main clinical and demographic factors between patients and controls (Supplementary Table 1).

### VIM location and extension

No differences were detected in the VIM volume between ET/N+ and ET/N- (0.13±0.09 cc vs. 0.18±0.12 cc, p= 0.99). The shape analysis revealed that the VIM extension was different between the two groups. In particular, the VIM parcels edges were more anterior relative to PC (2.8 ± 0.8 mm, p=0.04) and medial relative to midline (3.2 ± 2.6 mm, p=0.03) in ET/N+ than ET/N- cohorts (Figure 1a).

**Figure 1.**
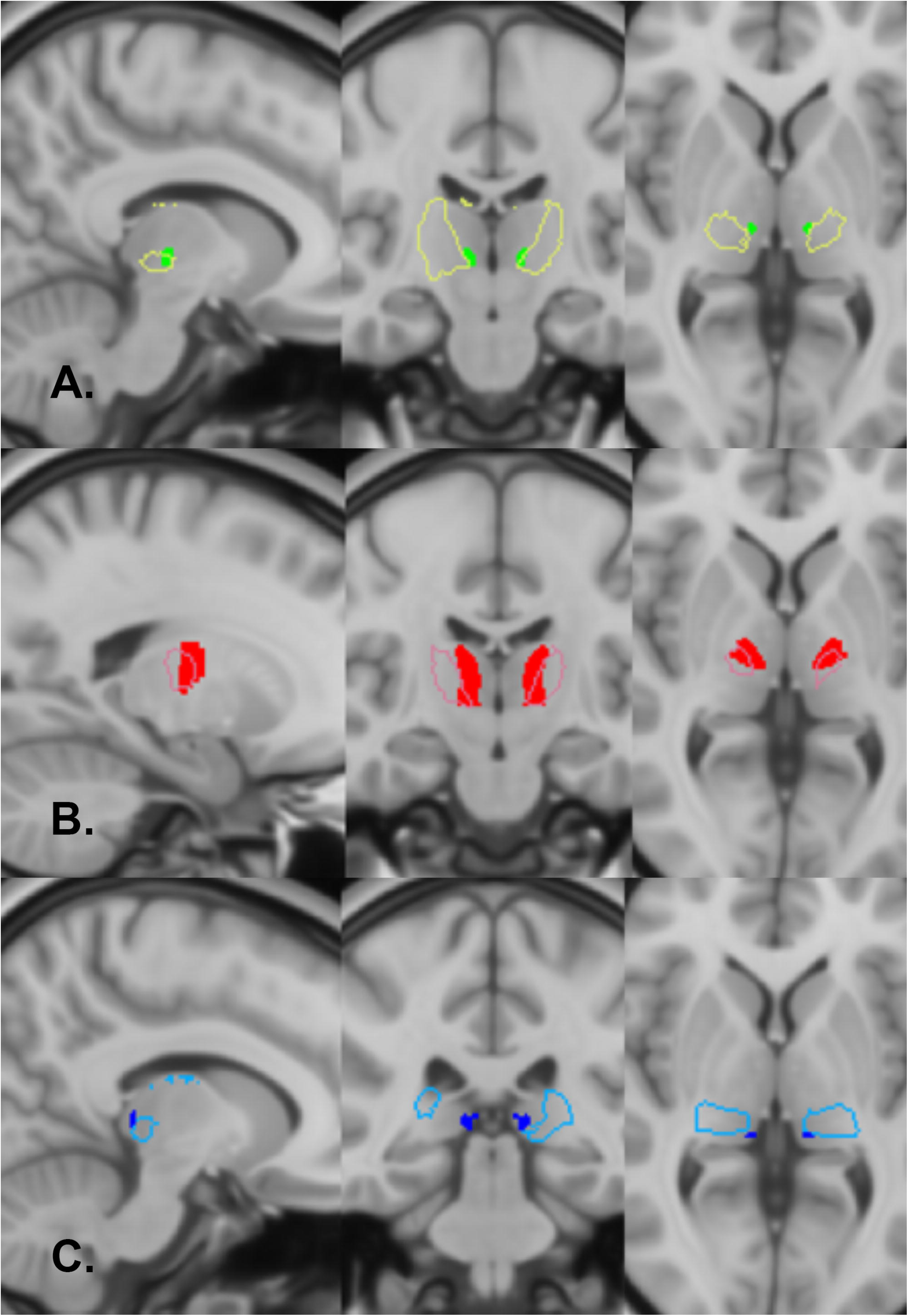
Shape analysis of the ventral thalamic nuclei revealed VIM (A) to be significantly more anterior and medial in the ET/N+ cohort than ET/N-. Similarly, VOA/VOP (B) was significantly more lateral and anterior, and Vc (C) more medial and posterior. The sagittal, coronal, and axial projections are shown with the boundaries of the grouped nuclei outlined with a line separately for VIM (yellow-green), VOA/VOP (red), and Vc (blue). The voxels significantly different between the ET/N+ and ET/N- cohorts are shown in solid colors (green: VIM, red: VOA/VOP, blue: Vc).

VC was found to be more medial (3.4 ±4.2 mm, p=0.04) and posterior (2.3 ± 5.1 mm, p=0.03) in ET/N+ than ET/N-, while the VOA/VOP parcel was more lateral (2.7 ± 1.8 mm, p=0.01) and anterior (3.7 ± 3.1 mm, p=0.03) (Figure 1b-c).

### Surgery-related factors

While all the electrode tips were located within the VIM, there was a significant difference in the mediolateral coordinates between ET/N+ and ET/N- (p=0.019). For the ET/N+ cohort, the mean coordinates of the electrode tips in the Montreal Neurological Institute template were: X = ±11.7 (SD 1.5) Y = −16.7 (SD 0.9), Z = −1.4 (SD 0.2). The respective coordinates for the ET/N- cohort were: X = ±15.4 (SD 1.5), Y = −15.7(SD 0.5), Z = −1 (SD 0.4). There were no significant differences in the number of passes (p= 0.21) or the Vim length (p= 0.36) identified with intraoperative electrophysiology. All patients in both groups achieved >50% intraoperative tremor reduction. The threshold for persistent paresthesias was significantly lower in the ET/N+ cohort (mean threshold for persistent paresthesia, ET/N+: 1.83 mA (SD 0.44), ET/N-: 2.84 mA (SD 1.38), p= 0.012).

### VTA analysis

The VTA volume was similar between ET/N+ and ET/N- (0.14±0.02 cc vs. 0.13±0.06 cc; p= 0.88), but more posterior (2.5 ±1.9 mm), dorsal (2 ±1.6 mm), and medial (1±1.8 mm) in ET/N+ than ET/N- (p < 0.04) (Figure 2). Also, the VTA coverage of the VIM was lower in ET/N+ than ET/N- (32±33% vs. 48±18%, p= 0.049). No differences were observed in the VTA volume and location between initial and 12-months programming (p= 0.99 and 0.76, respectively).

**Figure 2.**
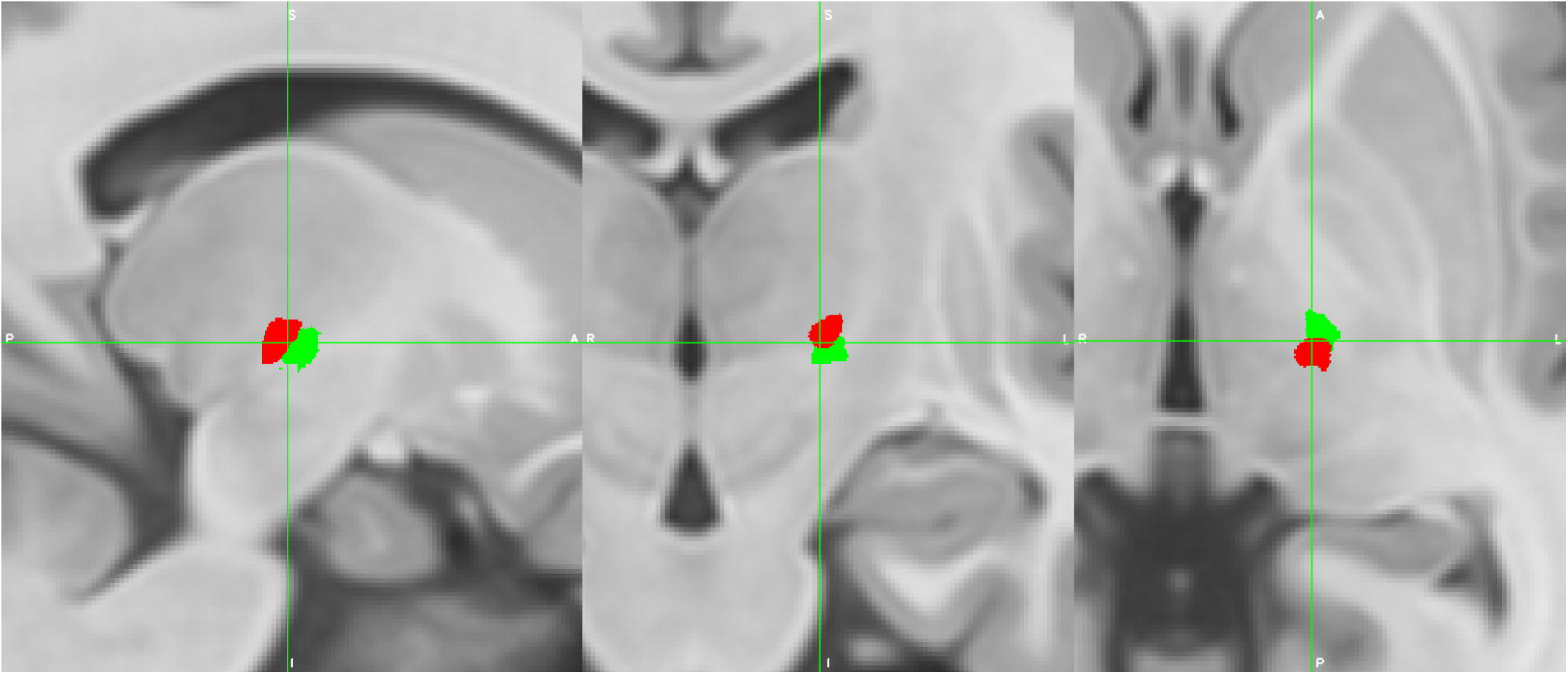
The volume of tissue activated was significantly different in the ET/N+ (red) and ET/N- (green) cohorts. The 75% overlap of the grouped stimulation volumes were significantly more posterior, dorsal, and medial in the ET/N+ cohort.

### Clinical outcomes and associated target coverage

The prevalence of suboptimal outcomes was higher in ET/N+ than ET/N- (13 out of 18 (72.2%) vs. 4 out of 18 (22.2%); χ2= 11.8, p= 0.0006), with a tremor improvement of 13.4%±11.4% (ET/N+) vs. 58.0%±13.4% (ET/N-) at 6 months (p= 2.125e-10) and 25.0%±13.0% (ET/N+) vs. 72.2%±10.8% (ET/N-) at 12 months (p= 1.52e-9) (Figure 3). At 12 months, tremor medications were increased or unchanged more commonly in ET/N+ (12 out of 18) than ET/N- (5 out of 18) (Chi-squared test, χ2= 4.65, p= 030).

**Figure 3.**
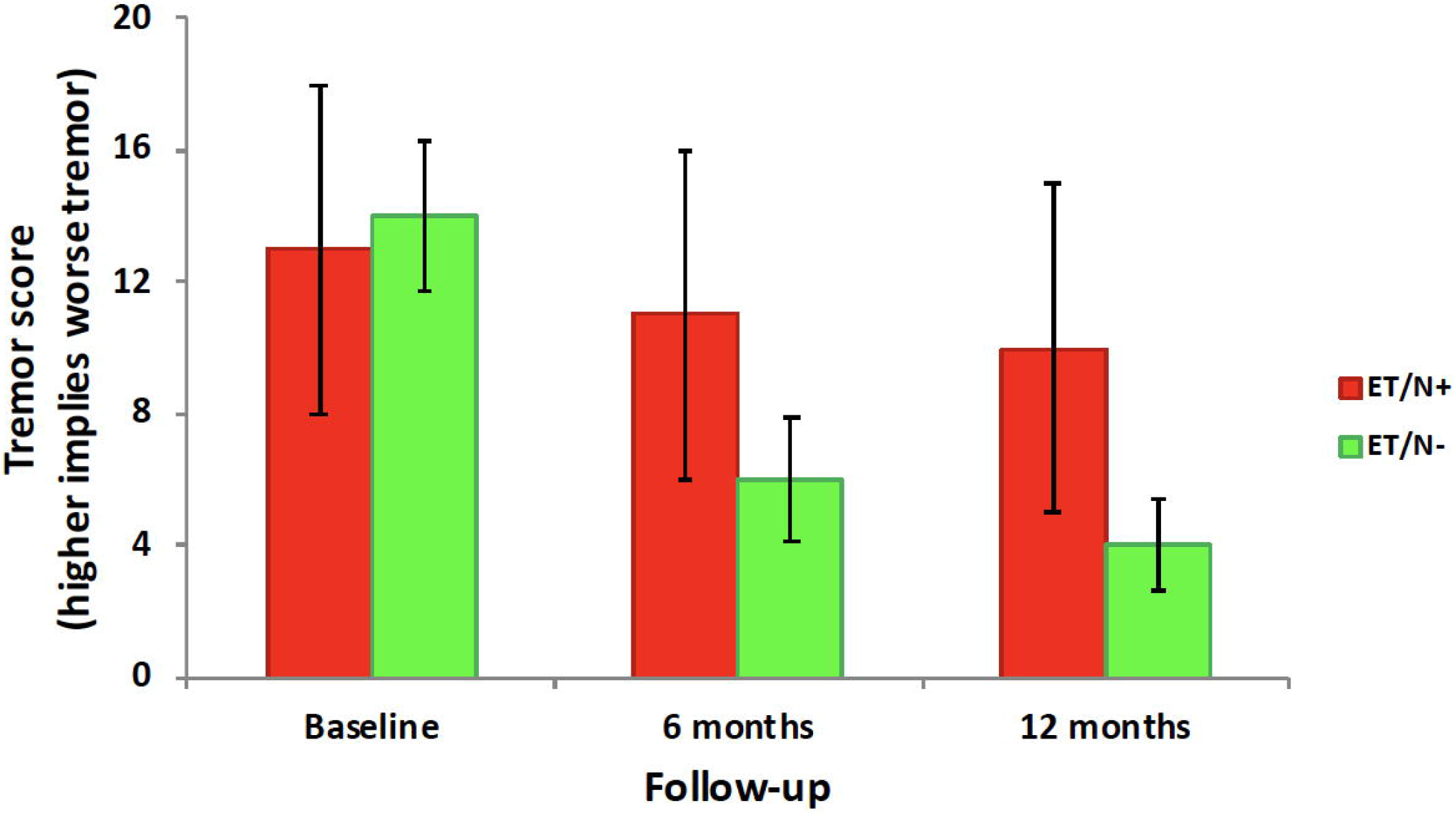
Tremor reduction was significantly lower in the ET/N+ (red) than ET/N- (green). Bar chart comparing mean tremor scores and standard deviations at baseline, 6 months, and 12 months.

There was an association between the volume of VIM covered by VTA and clinical outcomes. In ET/N-, a VIM coverage equal or greater than 95% was associated with better tremor outcomes at the 12-month follow-up (11 out of 12 patients with >50% tremor reduction) (Figure 4). In contrast, with a VIM coverage of 32%±33%, 13 out of 18 ET/N+ patients had <50% tremor reduction (Figure 5).

**Figure 4.**
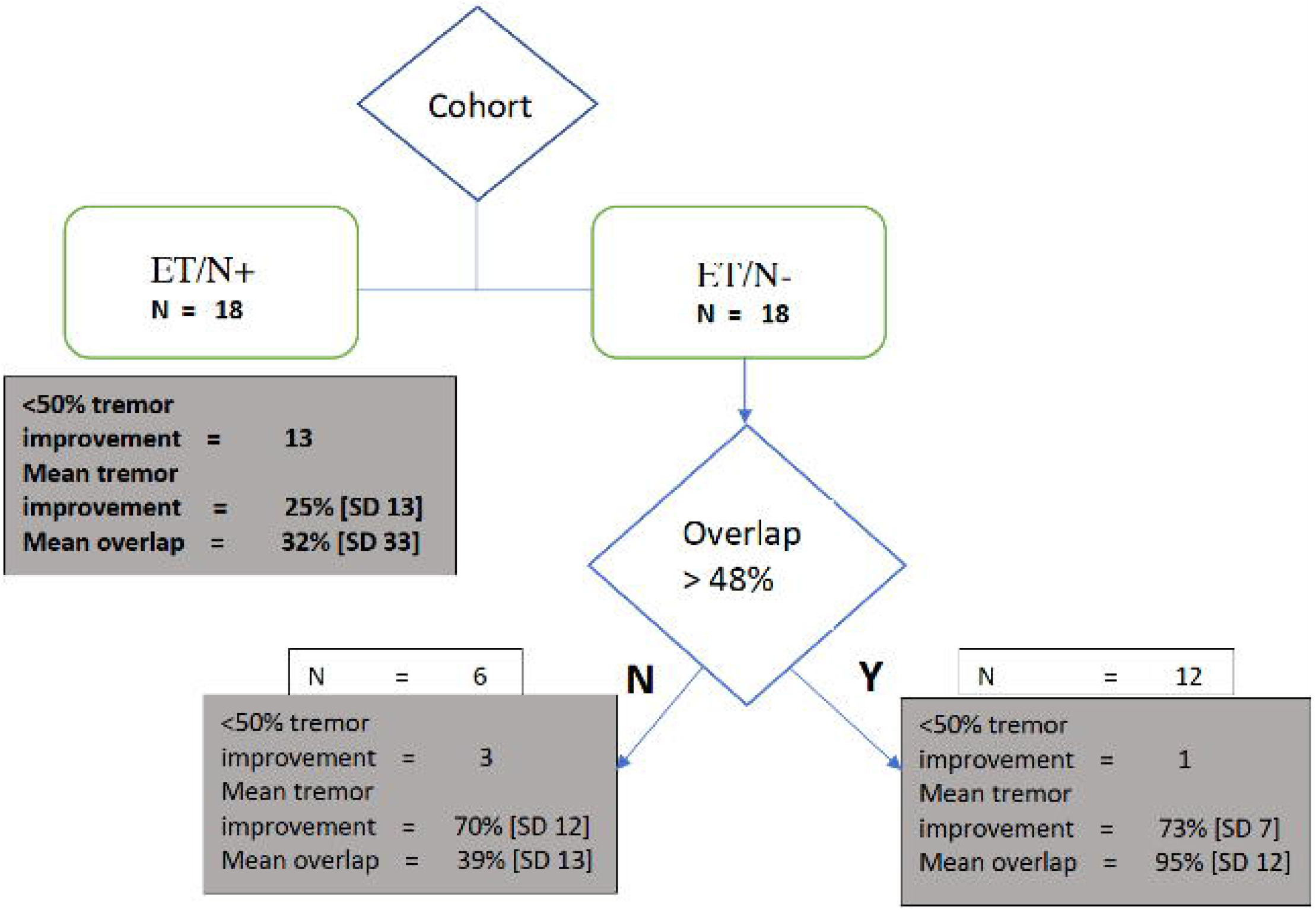
The overlap between VIM and tissue volume activated (VTA) by DBS was significantly associated with tremor outcomes. The recursive partitioning analysis results are shown. The patient cohort (ET/N+ or ET/N-) was the first node indicating significantly worse outcomes and low overlap between VIM and VTA in the ET/N+ cohort. For the ET/N- cohort, the best tremor outcomes were observed in patients with >48% overlap between VIM and VTA.

**Figure 5.**
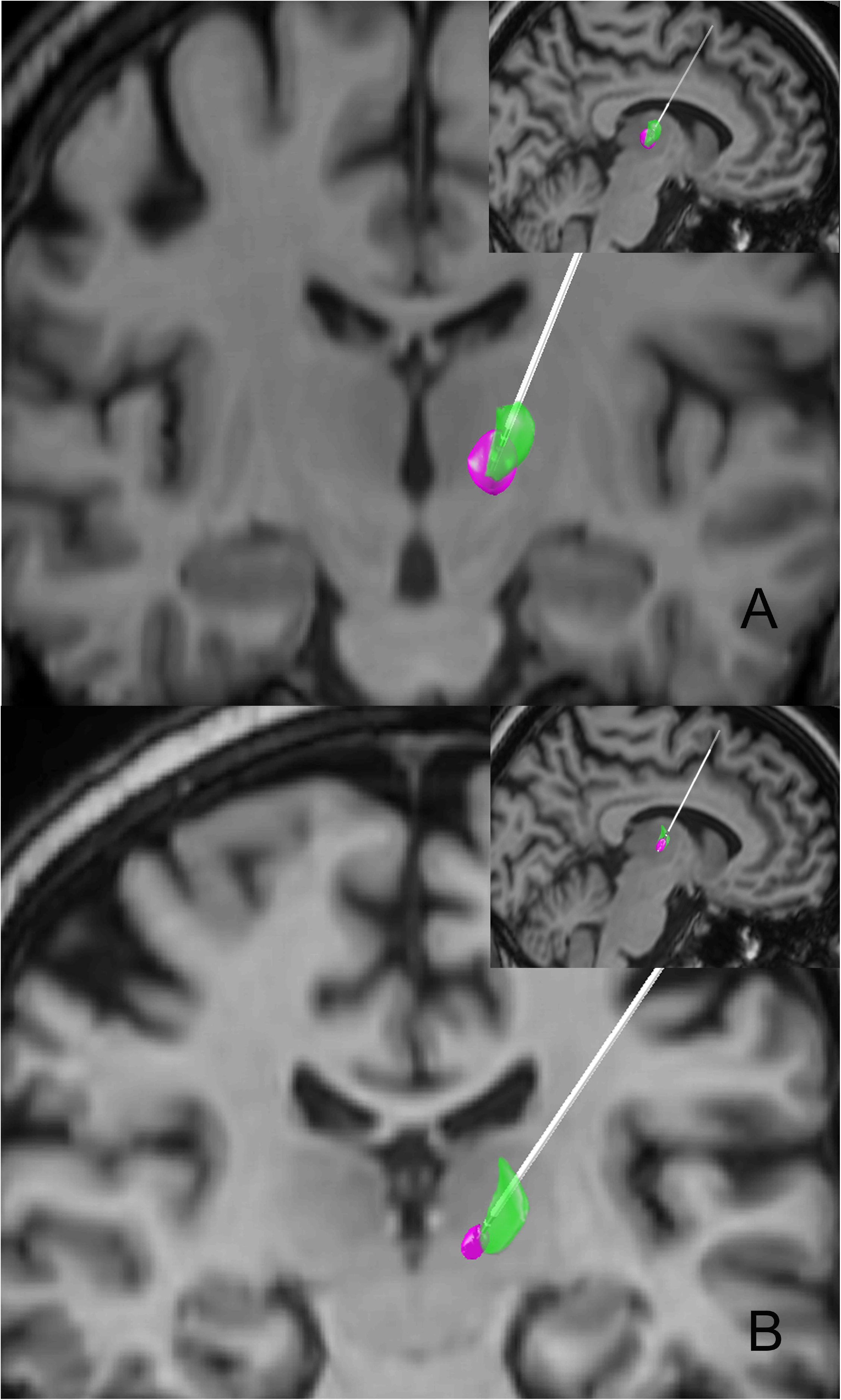
Representative images from two patients in the ET/N- (A) and ET/N+ (B) cohorts highlighting the differences in the overlap between VIM (green) and DBS VTA (purple). Coronal and sagittal (inset) projections show significantly higher overlap in the ET/N- than ET/N+ patient despite the electrode tip being in the VIM.

## Discussion

Using dMRI, we found that the location and extension of the VIM were significantly different in ET/N+ vs. ET/N- patients treated with VIM-DBS. The VIM was more anterior and medial, VC was more medial and posterior, and VOA/VOP complex was more lateral and anterior in ET/N+ than ET/N-, suggesting a thalamic reorganization of these structures in ET patients with comorbid neuropathy (disease-related factors). Despite similar surgery-related factors, the tremor outcomes at 12 months were suboptimal in ET/N+, partly due to increased programming difficulties reflected by a lower VIM target coverage and a lower threshold for persistent paresthesias.

Suboptimal outcomes were previously reported after DBS in patients with neuropathic tremor ^16, 31^. However, the tremor outcomes in patients with co-morbid PNP have not been fully characterized. Using a matched case-control design, we found that tremor outcomes were worse after VIM DBS in ET/N+ vs. ET/N-. Interestingly, no significant differences were observed in the baseline tremor scores between the two cohorts, confirming that a different baseline tremor severity did not bias the observed differences. Our results suggest that PNP was associated with significant differences in the location and extension of VIM and other ventral thalamic nuclei (VC, VOA/VOP), and suggest the possibility of a reorganization of these central nervous system structures in ET patients with comorbid neuropathy. We also showed that this thalamic reorganization presented with difficulties in postoperative programming, as indicated by a lower target coverage and a lower threshold for persistent paresthesia. Overall, these factors may potentially explain the reduced effectiveness of VIM-DBS in ET patients with comorbid neuropathy. Still, other disease-related factors may also affect the VIM-DBS outcomes. For example, it is well-known that PNP can worsen the clinical severity of ET ^32^ and potentially complicate its presentation due to sensory ataxia.^33^ Another possibility is that, besides anatomical reorganization, PNP may also lead to functional re-organization of VIM and other central nodes in the tremor network. ^13, 34, 35^ For example, previous studies suggested that PNP might lead to a reorganization of the primary sensorimotor cortex and cerebellum, which are modulated by VIM-DBS. ^36 17^

We report that VIM DBS surgery is less effective in ET/N+ patients. This finding can help guide the informed consent process while reviewing the risks and benefits and managing expectations after surgery. Future studies will need to test whether improving stereotactic VIM targeting and post-surgical DBS programming can salvage early VIM-DBS failure (within one year of implantation). Specifically, research should focus on testing tractography targeting combined with directional DBS electrodes in ET/N+ patients ^22, 23, 37^. In parallel, the connectivity-based selection of stimulation parameters might also prove useful in evaluating the thalamocortical connectivity to optimize the post-surgical programming ^38^. This approach is based on anatomically shaping the VTA model with directional DBS systems to optimized target coverage by preferentially steering the electrical field to obtain a better account for any shifts in VIM’s location and shape and improve the therapeutic window at a lower current ^39, 40^.

Some limitations should be acknowledged. First, we did not characterize the PNP severity with impairment scores to determine whether the severity of neuropathy predicted outcomes after DBS. Second, the number of patients included in the ET/N+ and ET/N- cohorts was inevitably limited by the rare prevalence of co-morbid PNP in ET patients. Third, the retrospective analysis of data and the lack of a-priori trial design introduce some methodological biases that temper the strength of our results. For example, we determined the PNP status by relying on historical records documenting the diagnosis, clinical testing, and review of previous electro-diagnostic studies, but did not perform additional diagnostic examinations to establish the severity of PNP at the time of surgery. Still, the consistent use of validated clinical scales with excellent inter-rater reliability such as the Fahn–Tolosa– Marin tremor rating scale partly limit these concerns ^41^. Finally, we cannot test whether the effects of neuropathy of thalamic reorganization are reproducible in different types of PNPs or independent of the ET diagnosis. Future studies will need to differentiate these factors by studying different PNP subgroups of patients and testing a cohort of PNP patients without ET.

Considering these limitations, this study’s results confirm the suboptimal outcomes of VIM-DBS in ET/N+ patients and provide strong evidence in support of structural reorganization of the thalamic anatomical structures involved in the therapeutic modulation of tremor in patients with PNP.

FS: 1. Research project: A. Organization, C. Execution; 2. Statistical Analysis: A. Design, B. Execution; 3. Manuscript Preparation: A. Writing of the first draft, B. Review and Critique.

VN: 1. Research project: A. Execution; 2. Statistical Analysis: A. Execution; 3. Manuscript Preparation: A. Writing of the first draft.

BC: 1. Research project: A. Organization; 2. Statistical Analysis: A. Review and Critique; 3. Manuscript Preparation: A. Review and Critique.

AM: 1. Research project: A. Organization; 2. Statistical Analysis: A. Review and Critique; 3. Manuscript Preparation: A. Review and Critique.

VK: 1. Research project: A. Conception, B. Organization, C. Execution; 2. Statistical Analysis: A. Design, B. Review and Critique; 3. Manuscript Preparation: A. Review and Critique.

## Data Availability

Due to confidentiality agreements, supporting data can only be made available to bona fide researchers subject to a non-disclosure agreement. Details of the data and how to request access are available from vibhor.krishna2osumc.edu-

## References

1. Louis ED, Rios E, Henchcliffe C. How are we doing with the treatment of essential tremor (ET)?: Persistence of patients with ET on medication: data from 528 patients in three settings. Eur J Neurol 2010;17(6):882–884.

2. Schneier FR, Barnes LF, Albert SM, Louis ED. Characteristics of social phobia among persons with essential tremor. J Clin Psychiatry 2001;62(5):367–372.

3. Louis ED, Machado DG. Tremor-related quality of life: A comparison of essential tremor vs. Parkinson’s disease patients. Parkinsonism Relat Disord 2015;21(7):729–735.

4. Findley LJ, Cleeves L, Calzetti S. Primidone in essential tremor of the hands and head: a double blind controlled clinical study. J Neurol Neurosurg Psychiatry 1985;48(9):911–915.

5. Koller WC. Long-acting propranolol in essential tremor. Neurology 1985;35(1):108–110.

6. O’Brien MD, Upton AR, Toseland PA. Benign familial tremor treated with primidone. Br Med J (Clin Res Ed) 1981;282(6259):178–180.

7. Benabid AL, Pollak P, Gervason C, et al. Long-term suppression of tremor by chronic stimulation of the ventral intermediate thalamic nucleus. Lancet 1991;337(8738):403–406.

8. Deuschl G, Raethjen J, Hellriegel H, Elble R. Treatment of patients with essential tremor. The Lancet Neurology 2011;10(2):148–161.

9. Benabid AL, Pollak P, Gao D, et al. Chronic electrical stimulation of the ventralis intermedius nucleus of the thalamus as a treatment of movement disorders. J Neurosurg 1996;84(2):203–214.

10. Pilitsis JG, Metman LV, Toleikis JR, Hughes LE, Sani SB, Bakay RA. Factors involved in long-term efficacy of deep brain stimulation of the thalamus for essential tremor. J Neurosurg 2008;109(4):640–646.

11. Papavassiliou E, Rau G, Heath S, et al. Thalamic Deep Brain Stimulation for Essential Tremor: Relation of Lead Location to Outcome. Neurosurgery 2004;54(5):1120–1130.

12. Sandoe C, Krishna V, Basha D, et al. Predictors of deep brain stimulation outcome in tremor patients. Brain Stimul 2018;11(3):592–599.

13. Favilla CG, Ullman D, Wagle Shukla A, Foote KD, Jacobson CEt, Okun MS. Worsening essential tremor following deep brain stimulation: disease progression versus tolerance. Brain 2012;135(Pt 5):1455–1462.

14. Handforth A, Parker GA. Conditions Associated with Essential Tremor in Veterans: A Potential Role for Chronic Stress. Tremor Other Hyperkinet Mov (N Y) 2018;8:517.

15. Cabanes-Martinez L, Del Alamo de Pedro M, de Blas Beorlegui G, Bailly-Bailliere IR. Long-Term Effective Thalamic Deep Brain Stimulation for Neuropathic Tremor in Two Patients with Charcot-Marie-Tooth Disease. Stereotact Funct Neurosurg 2017;95(2):102–106.

16. Patel N, Ondo W, Jimenez-Shahed J. Habituation and rebound to thalamic deep brain stimulation in long-term management of tremor associated with demyelinating neuropathy. Int J Neurosci 2014;124(12):919–925.

17. Ruzicka E, Jech R, Zarubova K, Roth J, Urgosik D. VIM thalamic stimulation for tremor in a patient with IgM paraproteinaemic demyelinating neuropathy. Mov Disord 2003;18(10):1192–1195.

18. Fasano A, Helmich RC. Tremor habituation to deep brain stimulation: Underlying mechanisms and solutions. Mov Disord 2019;34(12):1761–1773.

19. England J, Gronseth G, Franklin G, et al. Distal symmetric polyneuropathy: a definition for clinical research: report of the American Academy of Neurology, the American Association of Electrodiagnostic Medicine, and the American Academy of Physical Medicine and Rehabilitation. 2005;64(2):199–207.

20. Elble RJ. Diagnostic criteria for essential tremor and differential diagnosis. Neurology 2000;54(11 Suppl 4):S2–6.

21. Shahani BT. Tremor in peripheral neuropathy. London: Palgrave Macmillan, 1984.

22. Sammartino F, Krishna V, King NKK, et al. Tractography-Based Ventral Intermediate Nucleus Targeting: Novel Methodology and Intraoperative Validation. Mov Disord 2016;31(8):1217–1225.

23. King NKK, Krishna V, Basha D, et al. Microelectrode recording findings within the tractography-defined ventral intermediate nucleus. J Neurosurg 2017;126(5):1669–1675.

24. Behrens TE, Johansen-Berg H, Woolrich MW, et al. Non-invasive mapping of connections between human thalamus and cortex using diffusion imaging. Nat Neurosci 2003;6(7):750–757.

25. Horn A, Kuhn AA. Lead-DBS: a toolbox for deep brain stimulation electrode localizations and visualizations. Neuroimage 2015;107:127–135.

26. Krishna V, King NK, Sammartino F, et al. Anterior Nucleus Deep Brain Stimulation for Refractory Epilepsy: Insights Into Patterns of Seizure Control and Efficacious Target. Neurosurgery 2016;78(6):802–811.

27. Stacy MA, Elble RJ, Ondo WG, Wu SC, Hulihan J. Assessment of interrater and intrarater reliability of the Fahn-Tolosa-Marin Tremor Rating Scale in essential tremor. Mov Disord 2007;22(6):833–838.

28. Patenaude B, Smith SM, Kennedy DN, Jenkinson MJN. A Bayesian model of shape and appearance for subcortical brain segmentation. 2011;56(3):907–922.

29. Therneau TM, Atkinson EJ. An introduction to recursive partitioning using the RPART routines. Technical Report 61. URL http://www.mayo.edu/hsr/techrpt/61.pdf; 1997.

30. Peto R, Pike M, Armitage P, et al. Design and analysis of randomized clinical trials requiring prolonged observation of each patient. II. analysis and examples. 1977;35(1):1–39.

31. Blomstedt P, Fytagoridis A, Tisch S. Deep brain stimulation of the posterior subthalamic area in the treatment of tremor. Acta Neurochir (Wien) 2009;151(1):31–36.

32. Ahlskog MC, Kumar N, Mauermann ML, Klein CJ. IgM-monoclonal gammopathy neuropathy and tremor: a first epidemiologic case control study. Parkinsonism Relat Disord 2012;18(6):748–752.

33. Breit S, Wachter T, Schols L, et al. Effective thalamic deep brain stimulation for neuropathic tremor in a patient with severe demyelinating neuropathy. J Neurol Neurosurg Psychiatry 2009;80(2):235–236.

34. Louis ED, Faust PL, Vonsattel JP, et al. Neuropathological changes in essential tremor: 33 cases compared with 21 controls. Brain 2007;130(Pt 12):3297–3307.

35. Louis ED, Faust PL, Vonsattel JP, et al. Older onset essential tremor: More rapid progression and more degenerative pathology. Mov Disord 2009;24(11):1606–1612.

36. Rezai AR, Lozano AM, Crawley AP, et al. Thalamic stimulation and functional magnetic resonance imaging: localization of cortical and subcortical activation with implanted electrodes. Technical note. J Neurosurg 1999;90(3):583–590.

37. Krishna V, Sammartino F, Agrawal P, et al. Prospective Tractography-Based Targeting for Improved Safety of Focused Ultrasound Thalamotomy. Neurosurgery 2019;84(1):160–168.

38. Krishna V, Sammartino F, Rabbani Q, et al. Connectivity-based selection of optimal deep brain stimulation contacts: A feasibility study. Annals of Clinical and Translational Neurology 2019;6(7):1142–1150.

39. Artusi CA, Farooqi A, Romagnolo A, et al. Deep brain stimulation in uncommon tremor disorders: indications, targets, and programming. J Neurol 2018;265(11):2473–2493.

40. Merola A, Romagnolo A, Krishna V, et al. Current Directions in Deep Brain Stimulation for Parkinson’s Disease—Directing Current to Maximize Clinical Benefit. Neurology and Therapy 2020;9(1):25–41.

41. Stacy MA, Elble RJ, Ondo WG, Wu SC, Hulihan J, group TRSs. Assessment of interrater and intrarater reliability of the Fahn-Tolosa-Marin Tremor Rating Scale in essential tremor. Mov Disord 2007;22(6):833–838.

