## Supplementary material for "Thalamic organization in essential tremor patients with neuropathy: implication for functional neurosurgery": Table1

| **Variables** | | | **ET patients with neuropathy [n=23]** | **ET patients without neuropathy [n=23]** |
| --- | --- | --- | --- | --- |
| **Male gender** | | | 13 (56.5%) | 12 (52.2%) |
| **Age at surgery** | | | 68±7.8 (51-83) | 65±8.9 (54-85) |
| **Disease duration [years]** | | | 36±11.3 | 31±9.7 |
| **Family history of ET (n=41)** | | **Positive** | 13/20 [65%] | 14/21[66.7%] |
|  |  | **Negative** | 7/20 [35%] | 7/21 [33.3%] |
| **Tremor response to alcohol (n=30)** | | **Yes** | 8/14 (57.1%) | 9/16 (56.2%) |
|  |  | **No** | 6/14 (42.9%) | 7/16 (43.8%) |
| **Neuropathy** | **Diabetic neuropathy** | | 9 | NA |
|  | **Toxic/Drug/MS** | | 3 | NA |
|  | **Thyroid related neuropathy** | | 3 | NA |
|  | **Alcoholic neuropathy** | | 3 | NA |
|  | **Chemotherapy related neuropathy** | | 1 | NA |
|  | **Amyloid neuropathy** | | 1 | NA |
|  | **Vitamin B12 neuropathy** | | 2 | NA |
|  | **Paraproteinemia neuropathy** | | 1 | NA |
| **Tremor score [Mean, base line with range]** | | | 13±2.6 (9-18) | 14±2.3 (10-18) |
| **Tremor score [Mean, 6 month follow up with range]** | | | 11±2.9 (4-16) | 6±1.9 (0-8) |
| **Tremor score [Mean, 12 month follow up with range]** | | | 10±2.3 (4-14) | 4±1.4 (0-6) |
